# Rapid Epidemiological Analysis of Comorbidities and Treatments as risk factors for COVID-19 in Scotland (REACT-SCOT): a population-based case-control study

**DOI:** 10.1101/2020.05.28.20115394

**Authors:** Paul M McKeigue, Amanda Weir, Jen Bishop, Stuart J McGurnaghan, Sharon Kennedy, David McAllister, Chris Robertson, Rachael Wood, Nazir Lone, Janet Murray, Thomas M Caparrotta, Alison Smith-Palmer, David Goldberg, Jim McMenamin, Colin Ramsay, Sharon Hutchinson, Helen M Colhoun, On behalf of Public Health Scotland COVID-19 Health Protection Study Group

## Abstract

**Background:** The objectives of this study were to identify risk factors for severe COVID-19 and to lay the basis for risk stratification based on demographic data and health records.

**Methods and Findings:** The design was a matched case-control study. Severe COVID-19 was defined as either a positive nucleic acid test for SARS-CoV-2 in the national database followed by entry to a critical care unit or death within 28 days, or a death certificate with COVID-19 as underlying cause. Up to ten controls per case matched for sex, age and primary care practice were selected from the population register. All diagnostic codes from the past five years of hospitalisation records and all drug codes from prescriptions dispensed during the past nine months were extracted. Rate ratios for severe COVID-19 were estimated by conditional logistic regression.

There were 4272 severe cases. In a logistic regression using the age-sex distribution of the national population, the odds ratios for severe disease were 2.87 for a 10-year increase in age and 1.63 for male sex. In the case-control analysis, the strongest risk factor was residence in a care home, with rate ratio (95% CI) 21.4 (19.1, 23.9).

Univariate rate ratios (95% CIs) for conditions listed by public health agencies as conferring high risk were 2.75 (1.96, 3.88) for Type 1 diabetes, 1.60 (1.48, 1.74) for Type 2 diabetes, 1.49 (1.37, 1.61) for ischemic heart disease, 2.23 (2.08, 2.39) for other heart disease, 1.96 (1.83, 2.10) for chronic lower respiratory tract disease, 4.06 (3.15, 5.23) for chronic kidney disease, 5.4 (4.9, 5.8) for neurological disease, 3.61 (2.60, 5.00) for chronic liver disease and 2.66 (1.86, 3.79) for immune deficiency or suppression.

78% of cases and 52% of controls had at least one listed condition (NA of cases and NA of controls under age 40). Severe disease was associated with encashment of at least one prescription in the past nine months and with at least one hospital admission in the past five years [rate ratios 3.10 (2.59, 3.71)] and 2.75 (2.53, 2.99) respectively] even after adjusting for the listed conditions. In those without listed conditions significant associations with severe disease were seen across many hospital diagnoses and drug categories. Age and sex provided 2.58 bits of information for discrimination. A model based on demographic variables, listed conditions, hospital diagnoses and prescriptions provided an additional 1.25 bits (C-statistic 0.825). A limitation of this study is that records from primary care were not available.

**Conclusions:** Along with older age and male sex, severe COVID-19 is strongly associated with past medical history across all age groups. Many comorbidities beyond the risk conditions designated by public health agencies contribute to this. A risk classifier that uses all the information available in health records, rather than only a limited set of conditions, will more accurately discriminate between low-risk and high-risk individuals who may require shielding until the epidemic is over.

**Author summary:** Most people infected with the SARS-CoV-2 coronavirus do not become seriously ill. It is The risk of severe or fatal illness is higher in older than in younger people, and is higher in people with conditions such as asthma and diabetes than in people without these conditions. Using Scotland’s capability for linking electronic health records, we report the first systematic study of the relation of severe or fatal COVID-19 to pre-existing health conditions and other risk factors. We show that the strongest risk factor, apart from age, is residence in a care home. The conditions associated with increased risk include not only those already designated by public health agencies – asthma, diabetes, heart disease, disabling neurological disease, kidney disease – but many other diagnoses, associated with frailty and poor health. This lays a basis for constructing risk scores based on electronic health records that can be used to advise people at high risk of severe disease to shield themselves when there cases in their neighbourhood.

## Background

Case series from many countries have suggested that in those with severe COVID-19 the prevalence of diabetes and cardiovascular disease is higher than expected. For example in a large UK series the commonest co-morbidities were cardiac disease, diabetes, chronic pulmonary disease and asthma [1]. However there are also anecdotal reports of apparently healthy young persons succumbing to disease [2].

Quantification of the risk associated with characteristics and co-morbidities has been limited by the lack of comparisons with the background population [3–5]. Two recent studies in the UK have included population comparators and have reported associations of in hospital test positive persons and COVID-19 death in hospital with co-morbidities including diabetes, asthma and heart disease [6,7]. These studies have focused on conditions presumptively listed by public health agencies as increasing risk for COVID-19 based on case series data.

Here we examine the frequency of sociodemographic factors and these listed conditions in all people with severe COVID-19 disease in Scotland compared to matched controls from the general population. In those without listed conditions we report a systematic examination of the hospitalisation record and prescribing history in severe COVID-19 cases compared to controls. The objectives were to identify risk factors for severe COVID-19 and to lay the basis for risk stratification based on a predictive model.

## Methods

The study followed a pre-specified protocol. Modifications after study started were that we aligned the list of conditions of interest to be consistent with those designated as moderate risk conditions by public health agencies, and extended the list of drug classes under study to include all drugs.

### Case definition and selection of matched controls

All individuals testing positive for nucleic acid for SARS-CoV-2 were ascertained through the Electronic Communication of Surveillance in Scotland (ECOSS) database, which captures all virology testing in all NHS laboratories nationally. Linkage to other datasets was carried out using the Community Health Index (CHI) number, a unique identifier used in all care systems in Scotland. Admissions to critical care were obtained from the Scottish Intensive Care Society and Audit Group (SICSAG) database that captures admission to all critical care (intensive care or high dependency) units and has returned a daily census of patients in critical care from the beginning of the COVID-19 epidemic. Death registrations were obtained from linkage to the National Register of Scotland. Severe or fatal COVID-19 was defined by either (1) a positive nucleic acid test followed by entry to critical care or death within 28 days; or (2) a death certificate with COVID-19 as underlying cause. Using this definition ensures ascertainment of all severe cases even if they die without testing positive or entering critical care, whatever selection policies may have limited entry to critical care.

For each case, the CHI database was used to select up to ten controls matched for sex, one-year age band and registered with the same primary care practice, who were alive and resident in Scotland on the day as the first date that the case tested positive. For fatal cases who had not tested positive, the incident date was assigned as 14 days before death. To ensure that cases and controls were representative of the same population at risk, the 0.6% of cases who were recorded on the CHI database as no longer alive and resident in Scotland on the day that ECOSS recorded them as testing positive were also excluded. As this is an incidence density sampling design, it is possible and correct for an individual to appear in the dataset more than once, initially as a control and subsequently as a case.

For this analysis based on ascertainment of positive test results up to 6 June 2020, entry to critical care up to 14 June 2020 and deaths registered up to 14 June 2020 there were 4272 cases and 36948 controls. Among fatal cases, 94% of deaths were registered within 5 days.

### Demographic data

Residence in a care home was ascertained from the CHI database. Socioeconomic status was assigned as the Scottish Index of Multiple Deprivation (SIMD), an indicator based on postal code. For 74% of controls and 85% of cases self-assigned ethnicity, based on the categories used in the Census, had been recorded in Scottish Morbidity Records (SMR).

### Morbidity and drug prescribing

For all cases and controls, ICD-10 diagnostic codes were extracted from the last five years of hospital discharge records in the Scottish Morbidity Record (SMR01), excluding records of discharges less than 25 days before testing positive for SARS-CoV-2 and using all codes on the discharge. Diagnostic coding under ICD chapters 5 (Mental, Behavioural and Neurodevelopmental) and 15 (Pregnancy) is incomplete as most psychiatric and maternity unit returns are not captured in SMR01. British National Formulary (BNF) drug codes for dispensed prescriptions issued in primary care were extracted from the Scottish Prescribing Information System [8]. A cutoff date of 15 days before the incident date (date of testing positive for SARS-CoV-2, or 14 days before death for fatal cases without a positive test) was set, and prescriptions dispensed in a 240-day interval before this cutoff date were included. For this analysis prescription codes from BNF chapters 14 and above, comprising dressings, appliances, vaccines, anaesthesia and other preparations were grouped as “Other”.

We began by scoring a specific list of conditions that have been designated as risk conditions for COVID-19 by public health agencies [9]. A separate list of conditions designates “clinically extremely vulnerable” individuals who were advised to shield themselves completely since 24 March 2020: this list includes solid organ transplant recipients, people receiving chemotherapy for cancer, and people with cystic fibrosis or leukaemia. We did not separately tabulate these conditions as we expected these individuals to be underrepresented among cases if shielding was adequate.

The eight listed conditions were scored based on diagnostic codes in any hospital discharge record during the last five years, or encashed prescription of a drug for which the only indications are in that group of diagnostic codes. The R script included as supplementary material contains the derivations of these variables from ICD-10 codes and BNF drug codes. Diagnosed cases of diabetes were identified through linkage to the national diabetes register (SCI-Diabetes), with a clinical classification of diabetes type as Type 1, Type 2 or Other/Unknown.

Identifiable data were extracted by the Public Health Scotland CHI database and linkage team and de-identified before provision to the analysis team.

### Statistical methods

To estimate the relation of cumulative incidence and mortality from COVID-19 to age and sex, logistic regression models were fitted to the proportions of cases and non-cases in the Scottish population, using the estimated population of Scotland in mid-year 2019 which were available by one-year age group up to age 90 years. To allow for possible non-linearity of the relationship of the logit of risk to age, we also fitted generalized additive models, implemented in the R function gam::gam, with default smoothing function.

For the case-control study, all estimates of associations with severe COVID-19 were based on conditional logistic regression, implemented as Cox regression in the R function survival::clogit [10]. Among those cases and controls without any of the pre-defined conditions we then further examined associations of ICD-10 and BNF chapter with severe COVID-19. Restriction of cases and controls, for instance to exclude those with any listed condition, may generate strata that do not contain at least one case and at least one control, but these strata are ignored by the conditional logistic regression model as they do not contribute to the conditional likelihood. With incidence density sampling, the odds ratios in conditional logistic regression models are equivalent to rate ratios. Note that odds ratios in a matched case control study are based on the conditional likelihood and the unconditional odds ratios calculable from the frequencies of exposure in cases and controls will differ from these and should not be used [11].

Although matching on primary care practice will match to some extent for associated variables such as care home residence, socioeconomic disadvantage and prescribing practice, the effects of these variables are still estimated correctly by the conditional odds ratios but with less precision than in an unmatched study of the same size [11].

To construct risk prediction models, we used stepwise regression alternating between forward and backward steps to maximize the AIC, implemented in the R function stats::step. To evaluate the contribution of the listed conditions to risk prediction, and the incremental contribution of other information in hospitalisation and prescription records after assigning these conditions, predictive models were constructed from three sets of variables: a baseline set consisting only of demographic variables, a set that included indicator variables for each listed condition, and an extended set that included demographic, variables, indicator variables for listed conditions and indicator variables for hospital diagnoses in each ICD-10 chapter and prescriptions in each BNF chapter.

The performance of the risk prediction model in classifying cases versus non-cases of severe COVID-19 was examined by 10-fold cross-validation. We calculated the performance calculated over all test folds using the C-statistic but also using the “expected information for discrimination” Λ expressed in bits [12]. The use of bits (logarithms to base 2) to quantify information is standard in information theory: one bit can be defined as the quantity of information that halves the hypothesis space.

Although readers may be unfamiliar with the expected information for discrimination Λ, it has several properties that make it more useful than the C-statistic for quantifying increments in the performance of a risk prediction model [12]. A key advantage of using Λ is that contributions of independent predictors can be added. Thus in this study we can add the predictive information from a logistic model of age and sex in the general population to the predictive information provided by other risk factors from the case-control study matched for age and sex.

## Results

### Incidence and mortality from severe COVID-19 in the Scottish population

Figure 1 shows the relationships of incidence and mortality rates to age for each sex separately. The relationship of mortality to age is almost exactly linear on a logit scale, and the lines for male and female mortality are almost parallel. In models that included age and sex as covariates, the odds ratio associated with a 10-year increase in age was 2.87 for all severe disease and 3.7 for fatal disease. The odds ratio associated with male sex was 1.63 for all severe disease and 1.58 for fatal disease. For severe cases as defined in this study, the sex differential is narrow up to about age 50 but widens between ages 50 and 70 years. Thus at younger ages the ratio of critical care admissions to total fatalities is higher in women than in men, but that at later ages the ratio of critical admissions to total fatalities is higher in men.

**Fig 1.**
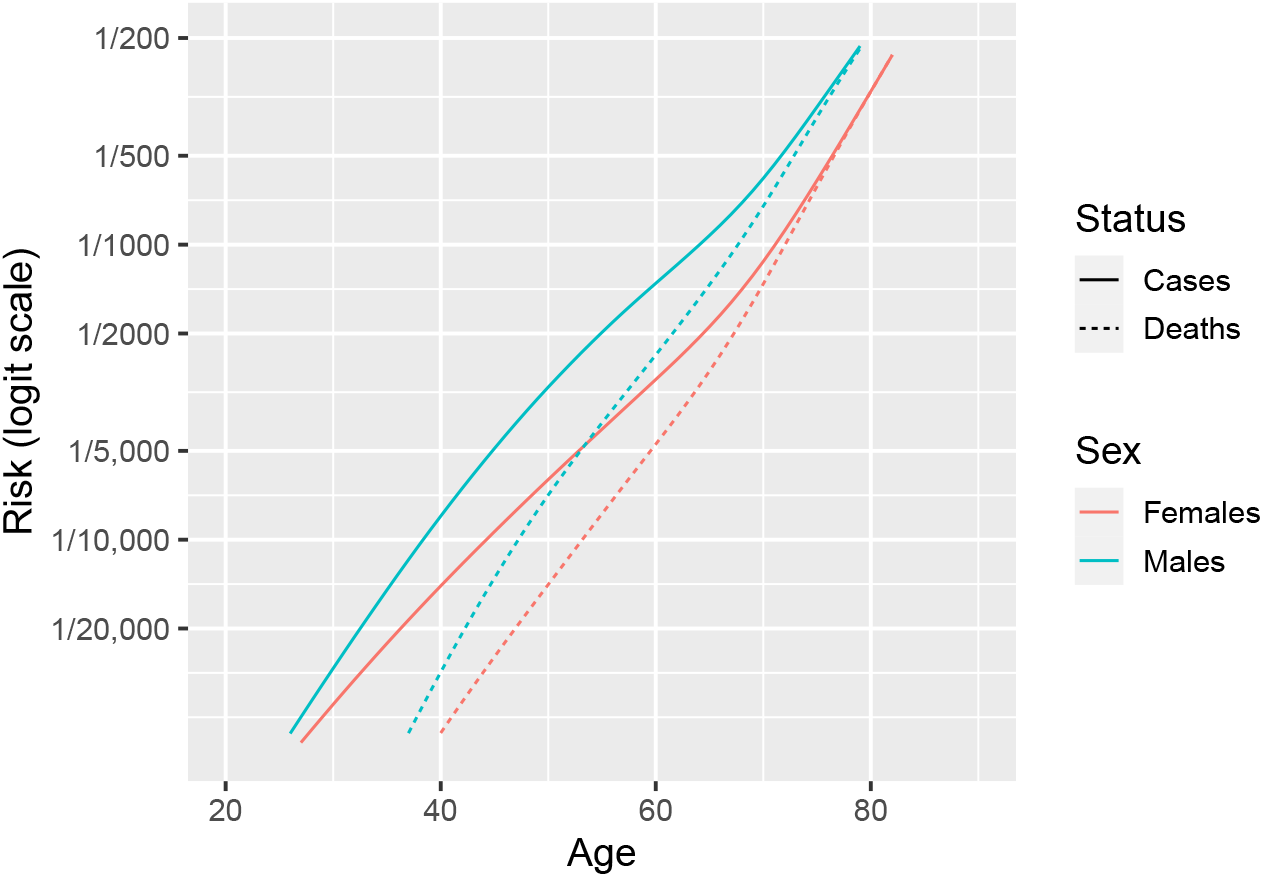
Incidence of severe and fatal COVID-19 in Scotland by age and sex: generalized additive models fitted to severe and fatal cases for males and females separately

### Risk factors

#### Sociodemographic factors

Table 1 shows univariate associations of demographic factors with severe disease. Residence in a care home was by far the strongest risk factor for severe disease. Higher risk of severe disease was also associated with socioeconomic deprivation. In the 85% of cases and 74% of controls in whom ethnicity had been recorded in the Scottish Morbidity Record, there were few non-White cases or controls and the confidence limits for the risk ratios for severe or fatal disease by ethnic group were wide.

**Table 1.** Univariate associations of severe disease with demographic factors

|  | Controls<br>(36948) | Cases<br>(4272) | Rate ratio (95%<br>CI) | *p*-value |
| --- | --- | --- | --- | --- |
| SIMD quintile |  |  |  |  |
| 1 - most deprived | 8559 (23%) | 1121 (26%) |  |  |
| 2 | 7956 (22%) | 935 (22%) | 0.86 (0.78, 0.95) | 0.003 |
| 3 | 6730 (18%) | 826 (19%) | 0.88 (0.79, 0.98) | 0.02 |
| 4 | 6558 (18%) | 773 (18%) | 0.81 (0.72, 0.90) | $2 \times 10^{-4}$ |
| 5 - least deprived | 7119 (19%) | 614 (14%) | 0.54 (0.48, 0.62) | $4 \times 10^{-21}$ |
| Care home | 2935 (8%) | 1894 (44%) | 21.4 (19.1, 23.9) | $8 \times 10^{-644}$ |
| Ethnicity based on Scottish Morbidity Record (27230 controls, 3648 cases) |  |  |  |  |
| White | 26908 (99%) | 3596 (99%) |  |  |
| South Asian | 145 (1%) | 27 (1%) | 1.26 (0.81, 1.97) | 0.3 |
| Black | 35 (0%) | 5 (0%) | 1.16 (0.44, 3.04) | 0.8 |
| Other | 142 (1%) | 20 (1%) | 1.01 (0.63, 1.65) | 1 |

#### Factors derived from hospitalisation and prescribing records

Prevalence of the listed conditions in cases and controls by age band is shown in Table 2. NA of the cases aged under 40 years had at least one listed condition, compared with only NA of the controls. In those aged 75+ years NA of the cases and NA of the controls had at least one listed condition. Among those aged under 40 years, NA of the cases and NA of the controls had either a hospital admission in the last five years or a dispensed prescription in the last 240 days. Differences in prescription rates between cases and controls narrowed with increasing age.

**Table 2.**
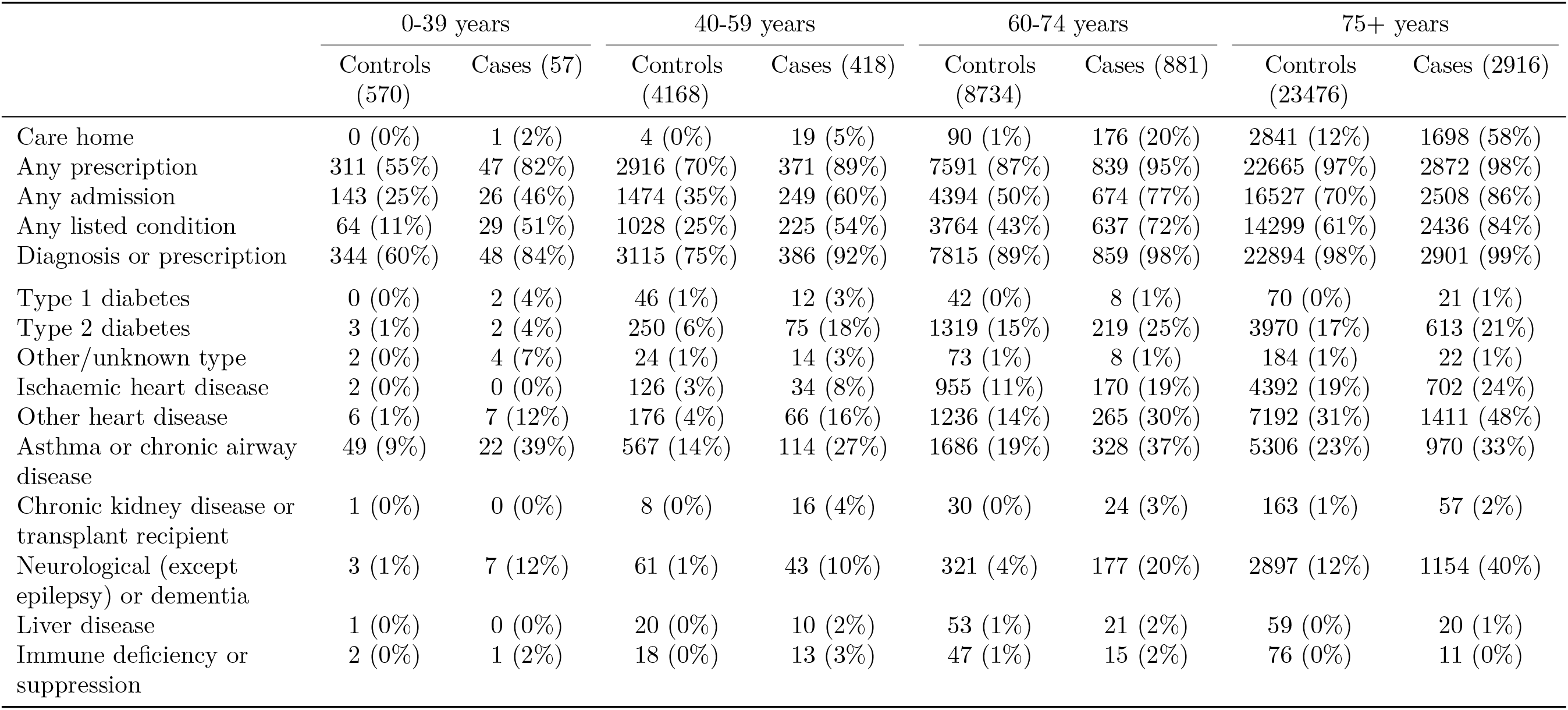
Frequencies of risk factors in cases and controls, by age group

|  | 0-39 years |  | 40-59 years |  | 60-74 years |  | 75+ years |  |
| --- | --- | --- | --- | --- | --- | --- | --- | --- |
|  | Controls<br>(570) | Cases (57) | Controls<br>(4168) | Cases (418) | Controls<br>(8734) | Cases (881) | Controls<br>(23476) | Cases (2916) |
| Care home | 0 (0%) | 1 (2%) | 4 (0%) | 19 (5%) | 90 (1%) | 176 (20%) | 2841 (12%) | 1698 (58%) |
| Any prescription | 311 (55%) | 47 (82%) | 2916 (70%) | 371 (89%) | 7591 (87%) | 839 (95%) | 22665 (97%) | 2872 (98%) |
| Any admission | 143 (25%) | 26 (46%) | 1474 (35%) | 249 (60%) | 4394 (50%) | 674 (77%) | 16527 (70%) | 2508 (86%) |
| Any listed condition | 64 (11%) | 29 (51%) | 1028 (25%) | 225 (54%) | 3764 (43%) | 637 (72%) | 14299 (61%) | 2436 (84%) |
| Diagnosis or prescription | 344 (60%) | 48 (84%) | 3115 (75%) | 386 (92%) | 7815 (89%) | 859 (98%) | 22894 (98%) | 2901 (99%) |
| Type 1 diabetes | 0 (0%) | 2 (4%) | 46 (1%) | 12 (3%) | 42 (0%) | 8 (1%) | 70 (0%) | 21 (1%) |
| Type 2 diabetes | 3 (1%) | 2 (4%) | 250 (6%) | 75 (18%) | 1319 (15%) | 219 (25%) | 3970 (17%) | 613 (21%) |
| Other/unknown type | 2 (0%) | 4 (7%) | 24 (1%) | 14 (3%) | 73 (1%) | 8 (1%) | 184 (1%) | 22 (1%) |
| Ischaemic heart disease | 2 (0%) | 0 (0%) | 126 (3%) | 34 (8%) | 955 (11%) | 170 (19%) | 4392 (19%) | 702 (24%) |
| Other heart disease | 6 (1%) | 7 (12%) | 176 (4%) | 66 (16%) | 1236 (14%) | 265 (30%) | 7192 (31%) | 1411 (48%) |
| Asthma or chronic airway disease | 49 (9%) | 22 (39%) | 567 (14%) | 114 (27%) | 1686 (19%) | 328 (37%) | 5306 (23%) | 970 (33%) |
| Chronic kidney disease or transplant recipient | 1 (0%) | 0 (0%) | 8 (0%) | 16 (4%) | 30 (0%) | 24 (3%) | 163 (1%) | 57 (2%) |
| Neurological (except epilepsy) or dementia | 3 (1%) | 7 (12%) | 61 (1%) | 43 (10%) | 321 (4%) | 177 (20%) | 2897 (12%) | 1154 (40%) |
| Liver disease | 1 (0%) | 0 (0%) | 20 (0%) | 10 (2%) | 53 (1%) | 21 (2%) | 59 (0%) | 20 (1%) |
| Immune deficiency or suppression | 2 (0%) | 1 (2%) | 18 (0%) | 13 (3%) | 47 (1%) | 15 (2%) | 76 (0%) | 11 (0%) |

Over all age groups, 3327 (78%) of severe cases and 19155 (52%) of controls had at least one of the listed conditions. As shown in Table 3, all the listed conditions were more frequent in cases than controls except for immune conditions in the 75+ age group. The rate ratio associated with type 1 diabetes was higher than that for type 2 diabetes. The rate ratio was 1.49 (1.37, 1.61) for ischemic heart disease compared to 2.23 (2.08, 2.39) for the broad category “other heart disease”. In multivariable analysis ischemic heart disease was not independently associated with severity whereas other heart disease remained strongly associated. In those without a listed condition 873 (92%) of the cases and 15052 (85%) of the controls had either a recent admission or a prescription. In those aged under 60 years without a listed condition, 184 (83%) of the cases and 2376 (65%) had either a recent admission or a prescription.

**Table 3.** Associations of severe disease with listed conditions over all age groups

|  |  | Controls<br>(36948) | Cases (4272) | Univariate |  | Multivariable |  |
| --- | --- | --- | --- | --- | --- | --- | --- |
|  |  |  |  | Rate ratio (95%<br>CI) | *p*-value | Rate ratio (95%<br>CI) | *p*-value |
| Care home | | 2935 (8%) | 1894 (44%) | 21.4 (19.1, 23.9) | $8 \times 10^{-644}$ | 14.7 (13.1, 16.6) | $1 \times 10^{-431}$ |
| Any prescription | | 33483 (91%) | 4129 (97%) | 3.10 (2.59, 3.71) | $8 \times 10^{-35}$ | 1.83 (1.51, 2.22) | $6 \times 10^{-10}$ |
| Any admission | | 22538 (61%) | 3457 (81%) | 2.75 (2.53, 2.99) | $2 \times 10^{-124}$ | 1.56 (1.41, 1.72) | $1 \times 10^{-18}$ |
| Type 1 diabetes | | 158 (0%) | 43 (1%) | 2.75 (1.96, 3.88) | $6 \times 10^{-9}$ | 1.56 (1.05, 2.32) | 0.03 |
| Type 2 diabetes | | 5542 (15%) | 909 (21%) | 1.60 (1.48, 1.74) | $8 \times 10^{-30}$ | 1.42 (1.29, 1.56) | $3 \times 10^{-13}$ |
| Other/unknown type | | 283 (1%) | 48 (1%) | 1.74 (1.28, 2.38) | $4 \times 10^{-4}$ | 1.58 (1.11, 2.27) | 0.01 |
| Ischaemic heart disease | | 5475 (15%) | 906 (21%) | 1.49 (1.37, 1.61) | $3 \times 10^{-21}$ | 1.08 (0.98, 1.20) | 0.1 |
| Other heart disease | | 8610 (23%) | 1749 (41%) | 2.23 (2.08, 2.39) | $4 \times 10^{-109}$ | 1.33 (1.22, 1.46) | $2 \times 10^{-10}$ |
| Asthma or chronic airway<br>disease | | 7608 (21%) | 1434 (34%) | 1.96 (1.83, 2.10) | $2 \times 10^{-78}$ | 1.54 (1.42, 1.68) | $7 \times 10^{-25}$ |
| Chronic kidney disease or<br>transplant recipient | | 202 (1%) | 97 (2%) | 4.06 (3.15, 5.23) | $3 \times 10^{-27}$ | 2.88 (2.13, 3.89) | $7 \times 10^{-12}$ |
| Neurological (except epilepsy) or<br>dementia | | 3282 (9%) | 1381 (32%) | 5.4 (4.9, 5.8) | $1 \times 10^{-354}$ | 2.00 (1.81, 2.21) | $2 \times 10^{-42}$ |
| Liver disease | | 133 (0%) | 51 (1%) | 3.61 (2.60, 5.00) | $2 \times 10^{-14}$ | 1.93 (1.32, 2.81) | $6 \times 10^{-4}$ |
| Immune deficiency or<br>suppression | | 143 (0%) | 40 (1%) | 2.66 (1.86, 3.79) | $7 \times 10^{-8}$ | 1.67 (1.10, 2.52) | 0.01 |

Supplementary Tables S2 to S4 examine these associations by age group, with the 0-39 and 40-59 year age bands combined. All listed conditions were associated with severe disease in each age band. In those aged under 60 years, the rate ratio was 3.70 (2.01, 6.79) for Type 1 diabetes and 3.70 (2.80, 4.90) for Type 2 diabetes. The multivariable analyses shown in Table 3 and S2 to S4 show that overall and in each age group group any admission to hospital in the past five years were strongly and independently associated with severe disease even after adjusting for care home residence and listed conditions. Dispensing of any prescription in the past year was associated with severe disease in multivariable analyses in the two younger age bands. Table 4 shows that in each age group the proportion of fatal cases who had not had either a hospital admission in the last five years or a dispensed prescription in the last year was very low.

**Table 4.** Proportions of fatal cases and matched controls without and with a dispensed prescription or hospital diagnosis, by age group

|  | Controls | Fatal cases |
| --- | --- | --- |
| **Age <60** |  |  |
| No prescription or diagnosis | 1305 (26%) | 15 (7%) |
| Prescription or diagnosis | 3696 (74%) | 197 (93%) |
| **Age 60-74** |  |  |
| No prescription or diagnosis | 929 (10%) | 12 (2%) |
| Prescription or diagnosis | 7994 (90%) | 680 (98%) |
| **Age 75+** |  |  |
| No prescription or diagnosis | 583 (2%) | 14 (0%) |
| Prescription or diagnosis | 22924 (98%) | 2871 (100%) |

#### Comparison of fatal and non-fatal cases

Supplementary Table S1 shows a breakdown of severe cases by test-positive status, entry to critical care, and fatal versus non-fatal outcome. Severe cases who entered critical care were much younger than severe cases never entering critical care. The majority of severe cases from residential care homes never entered critical care. Among fatal cases who did not enter critical care. the distribution of age and other risk factors was similar in those with and without a positive test result, except that the proportion of care home residents was higher among those without a positive test result. Among those entering critical care the median age and the proportion of males and prevalence or prior comorbidities were higher in fatal than in non-fatal cases.

#### Systematic analysis of diagnoses associated with severe disease

The association of severe COVID-19 with prior hospital admission was examined further by testing for association of hospitalisations at each ICD-10 chapter level with severe COVID-19, among those without any of the listed conditions. These results are shown in Supplementary Table S5. In univariate analyses, almost all ICD-10 chapters, with the exception of Chapters 7 (eye) Chapters 8 (ear) and Chapter 15 (pregnancy) were associated with increased risk of severe disease. In a multivariable analysis the strongest associations were with diagnoses in ICD chapters 4 (mental disorders) and 10 (respiratory). Supplementary Table S7 extracts univariate associations with ICD-10 subchapters in those without any listed conditions. This table is filtered to show only subchapters for which there are at least 50 cases and controls and the univariate p-value is <0.001. This shows that many subchapter diagnoses are associated with markedly higher risk of severe COVID-19.

#### Associations of prescribed drugs with severe disease

As shown in Table 3 and supplementary Tables S2 to S4, encashment of at least one prescription in the last year was associated with severe disease. The univariate rate ratio associated with this variable varies from 3.74 (2.79, 5.01) in those aged under 60 years to 2.30 (1.69, 3.14) in those aged 75 years and over. In a multivariable analysis adjusting for care home residence, any hospital admission and listed conditions, these rate ratios were reduced to 2.12 (1.55, 2.90) and 1.13 (0.80, 1.60) respectively.

To investigate this further, we partitioned the “Any prescription” variable into indicator variables for each chapter of the British National Formulary, in which drugs are grouped by broad indication, and restricted the analysis to those without one of the listed conditions. Table S6 shows these associations. In univariate analyses, prescriptions in almost all BNF chapters were associated with severe disease. In a multivariable analysis of all chapters, the strongest independent associations with severe disease were with prescriptions in chapters 1 (gastrointestinal), 4 (central nervous system), 5 (infections), 9 (nutrition and blood) and 14+ (other, mostly dressings and appliances).

#### Construction of a multivariable risk prediction model

The variables retained from the extended variable set (demographic variables, listed conditions, hospital diagnoses in each ICD-10 chapter, prescriptions in each BNF chapter) are shown in Table S8. Coefficients for specific conditions here should not be interpreted as effect estimates, as global variables for any hospital diagnosis and any listed condition have been included in the model. The predictive performance of the model chosen by stepwise regression was estimated by 10-fold cross-validation.

Observed and predicted case status were compared within each stratum over all test folds. Table 5 shows that using the extended set increased the C-statistic from 0.805 to 0.825 and the expected information for discrimination Λ from 1.09 bits to 1.25 bits.

**Table 5.** Prediction of severe COVID-19: cross-validation of models chosen by stepwise regression

| | Cases /<br>controls | Crude C-statistic | Adjusted<br>C-statistic | Crude $\Lambda$<br>(bits) | Adjusted<br>$\Lambda$ (bits) | Test log-<br>likelihood<br>(nats) |
| --- | --- | --- | --- | --- | --- | --- |
| Demographic<br>only | 4246 /<br>36719 | 0.789 | 0.768 | 0.97 | 0.86 | 0.0 |
| Demographic +<br>listed conditions | 4246 /<br>36719 | 0.826 | 0.805 | 1.19 | 1.09 | 414.3 |
| Extended<br>variable set | 4246 /<br>36719 | 0.839 | 0.825 | 1.33 | 1.25 | 670.1 |

Figure 2 shows the distributions in cases and controls of the weight of evidence favouring case over control status from the model based on the extended variable set with a footnote explaining how Λ is derived. This shows, as expected for a multifactorial classifier, that the distribution in controls is approximately Gaussian: there is no clear divide between high-risk and low-risk individuals of the same age and sex. In cases there is a bimodal distribution, where the second mode represents care home residents. Figure 3 shows the receiver operating characteristic curve with a footnote explaining its derivation from the distributions of the weights of evidence.

**Fig 2.**
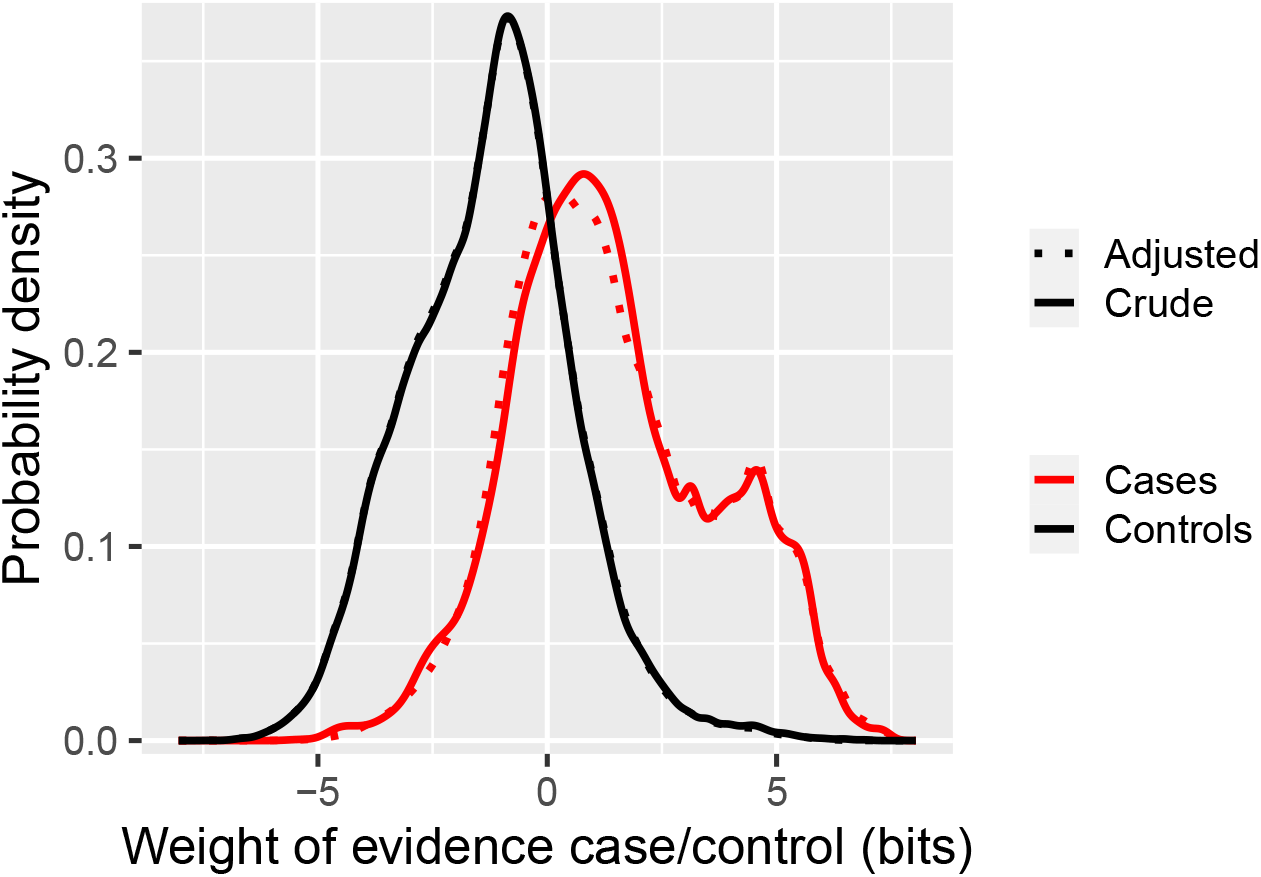
Cross-validation of model chosen by stepwise regression using extended variable set: class-conditional distributions of weight of evidence

[**Footnote for Figure 2** For each individual, the risk prediction model outputs the posterior probability of being a case, which can also be expressed as the posterior odds. Dividing the posterior odds by the prior odds gives the likelihood ratio favouring case over non-case status for an individual. The weight of evidence *W* is the logarithm of this ratio. The distributions of *W* in cases and controls in the test data are plotted in Figure 2. For a classifier, the further apart these curves are, the better the predictive performance. The expected information for discrimination Λ is the average of the mean of the distribution of *W* in cases and minus 1 times the mean of the distribution of *W* in controls. The distributions have been adjusted by taking a weighted average to make them mathematically consistent [12].]

**Fig 3.**
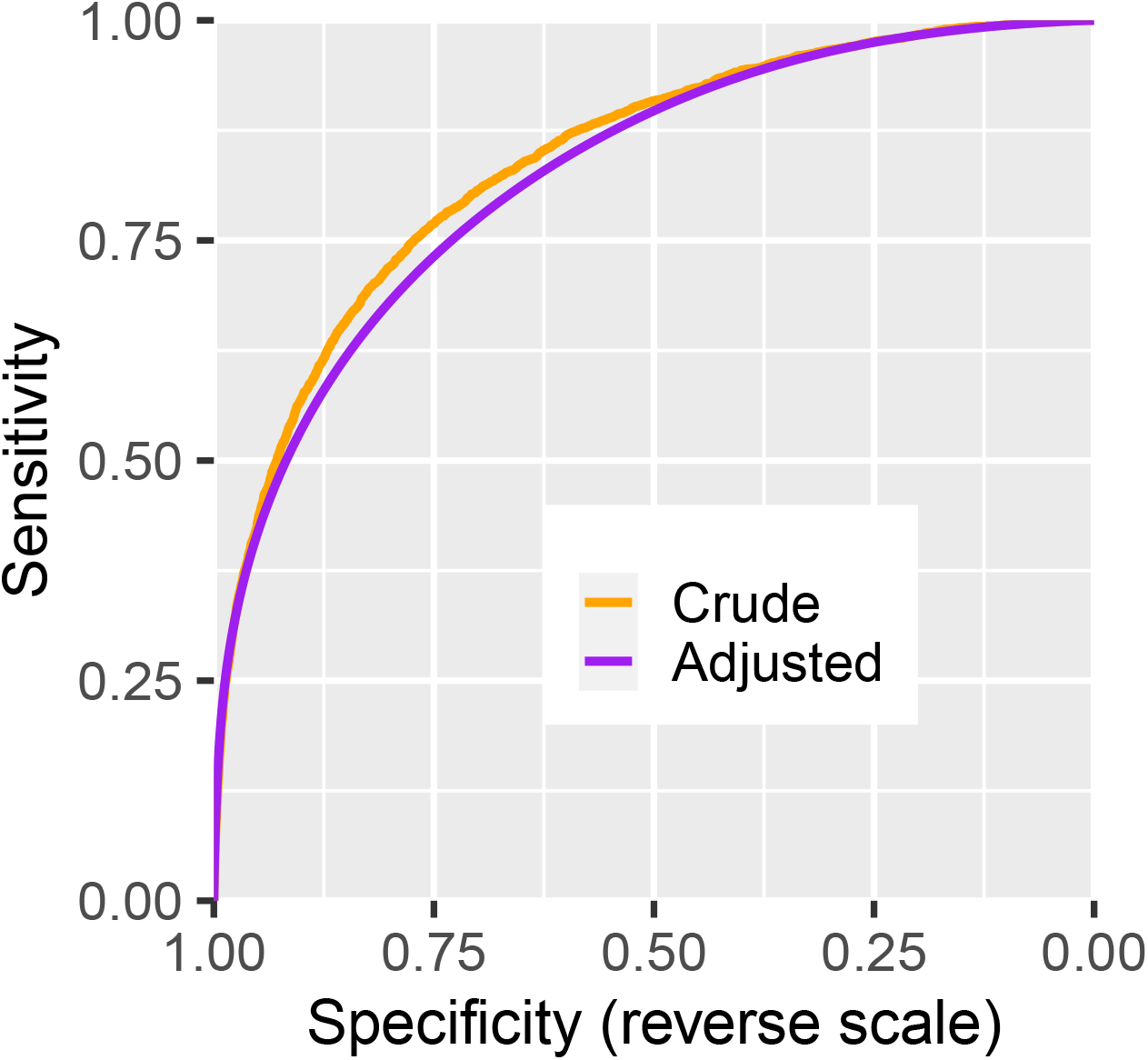
Cross-validation of model chosen by stepwise regression using extended variable set: receiver operating characteristic curve

[**Footnote for Figure 3** The receiver operator characteristic (ROC) curve is computed by calculating at each value of the risk score the sensitivity and specificity of a classifier that uses this value as the threshold for classifying cases and non-cases. Using the adjusted distributions from Figure 2 gives a curve that is concave downwards. The C-statistic is the area under this curve, computed as the probability of correctly classifying a case/noncase pair using the risk score, evaluated over all possible such pairs in the dataset.]

This estimate of 1.25 bits for the information conditional on age and sex obtained from the matched case-control study can be added to the information for discrimination 2.58 bits obtained from the logistic regression on age and sex in the population using age and sex to estimate the total information for discrimination of a risk classifier that would be obtained in the population as 3.83 bits.

## Discussion

### Sociodemographic factors

This analysis confirms that risk for severe COVID-19 is associated with increasing age, male sex and socioeconomic deprivation. The slope of the relationship of severe disease (on the scale of log odds) to age is less steep than the slope of the relationship of fatal disease to age. Residence in a care home was associated with a 21-fold increased rate of severe COVID-19 in this age matched analysis, reduced to 15-fold by adjustment for listed conditions. This excess risk is likely to reflect both the spread of the epidemic in care homes and residual confounding by frailty.

Although the assignment of ethnicity is incomplete and the prevalence of non-White ethnic groups in Scotland is low, we show the data as they give some indication of the likely upper bound of the absolute numbers of severe cases in South Asian and Black ethnic groups in Scotland. This is relevant because substantial elevations in risk of hospitalised and fatal COVID-19 have been reported elsewhere in the UK [6,7,13]. In the OpenSAFELY study risk ratios for fatal COVID-19 of 1.7 in those recorded as Black and and 1.6 in those recorded as Asian, in comparison with those recorded as White, persisted after adjustment for comorbidities and socioeconomic status. In Scotland however, although point estimates for rate ratios in those of South Asian and Black ethnicity compared to Whites are above 1, the confidence intervals are too wide for any definitive conclusions on ethnic differences.

### Co-morbidities

We have confirmed that the moderate risk conditions designated by the NHS and other agencies [9] are associated with increased risk of severe COVID-19. However the rate ratios associated with these conditions vary with age - for example the rate ratio associated with diabetes is higher at younger ages. The rate ratios of 2.8 for Type 1 diabetes and 1.6 for Type 2 diabetes are broadly similar to those reported in UK Biobank and in the OpenSAFELY studies. We confirm the higher risk with asthma and chronic lung disease and liver disease reported in these and earlier studies. An unexpected finding was that the risk associated with other forms of heart disease is higher than that associated with ischaemic heart disease. This category includes conditions such as atrial fibrillation, cardiomyopathies and heart failure. One of the highest rate ratios is that associated with chronic kidney disease. Prevention of nosocomial transmission in dialysis units may help to reduce this risk. Over all age groups, 78% of severe cases had at least one of these listed conditions. In this dataset it is not possible to examine adequately the risk associated with neoplasms, as we cannot separately identify those who were advised to shield themselves because they had active neoplasms of lymphoid or hematopoietic tissue or were receiving treatments that affect the immune system. We hope to explore this in a separate study that will include linkage to records of shielding advice. In patients without any listed conditions, further systematic evaluation of past hospitalisation history did not reveal a sparse set of underlying conditions; instead many diagnoses were associated with severe COVID-19.

Media reports of apparently healthy young people succumbing to severe COVID-19 have disseminated the message that all are at risk of disease whatever their age or health status. However we found that half of cases under 40 years had at least one of the listed conditions and among those who did not have one of these conditions, the proportions who had at least one prior hospitalisation or dispensed prescription were higher in cases than in controls. In all age groups, very few of the fatal cases had not had either a hospital admission in the past five years or a dispensed prescription in the past year.

A striking finding of this study was the association of severe COVID-19 with dispensing of at least one prescription in the past year, only partly explained by higher rates of prescribing among those with listed conditions. Partitioning of this association between BNF chapters, which represent broad indication-based drug classes, showed that prescription of drugs for the gastrointestinal and central nervous systems, together with nutritional supplements, contributed to this association. Although it is likely that most associations of severe COVID-19 with drug prescribing are attributable to the indications for which these drugs were prescribed, or to more diffuse frailty especially in older persons, causal effects of drugs or direct effects of polypharmacy on susceptibility cannot be ruled out. These associations are explored in a separate paper.

### Relevance to policy

As lockdown restrictions are eased, there is general agreement that vulnerable individuals will require shielding, even if the restart of the epidemic can be slowed or suppressed by mass testing, contact tracing and isolation of those who test positive. The “stratify and shield” policy option [14], in which high-risk individuals comprising up to 15% of the population are shielded for a defined period while the epidemic is allowed to run relatively quickly in low-risk individuals until population-level immunity is attained, depends critically on informative risk discrimination. So too does the similarly named “segment and shield” option [15] which has the opposite objective of keeping transmissions low. Although for this preliminary study we have not used the full repertoire of machine learning methods available for this type of problem, we have shown that a model based on health records provides 1.25 bits of information for discrimination conditional on age and sex. Adding this to the 2.58 bits provided by age and sex gives a total information for discrimination of 3.8 bits. \hl{We have shown elsewhere that this level of predictive performance would allow at least 80% of those at risk of severe or fatal disease to be allocated to a shielded group comprising no more than 15% of the population} [14]

As awareness grows of how risk varies between individuals, individuals will seek information about their own level of risk. A key implication of our results is that risk of severe or fatal disease is multifactorial. The rate ratio of 2.9 associated with a 10-year increase in age is stronger than the rate ratios associated with common diseases such as asthma or Type 2 diabetes that are listed as conditions associated with high risk. A corollary of this is that a crude classification based on assigning all persons with a listed condition to a group for whom shielding is recommended will have poor specificity, as one quarter of those aged 60-74 years in the population have at least one of the listed conditions we examined. It will also exclude many people at high risk because they have multiple risk factors each of small effect. A more meaningful way to score risk for an individual would be to use all available information to calculate a “COVID age” as the age at which the average risk for someone of the same sex in the population equates to the risk for the individual under study. Thus the rate ratio of 2.8 associated with Type 1 diabetes equates to an increase of 9.8 years in COVID age. In Scotland it is technically possible to use existing electronic health records to calculate a risk score for every individual in the population, though more work would be required to develop this as a basis for official advice and individual decisions.

### Methodological strengths and weaknesses

Most reports of disease associations with COVID-19 have been case series. There have been few reports based on evaluating these associations in the population through cohort or case-control studies. With this matched case control design using incidence density sampling, we have been able to estimate rate ratios conditional on age and sex. An unpublished analysis from England explored the association of similar set of risk conditions with in-hospital COVID-19 deaths, but did not systematically evaluate the rest of the medical record including prescription records. Although we have records of encashment of prescriptions, we do not at present have access to other primary care data, which would contain additional information on morbidity and measurements such as body mass index. A strength of our study however is that hospital discharge diagnoses are coded to ICD-10 by trained coders, in contrast to the coding systems used in primary care databases that do not map to recognized disease classifications. Associations with ethnicity and other sociodemographic factors are not necessarily generalizable from Scotland to other populations.

## Conclusion

This study confirms that risk of severe COVID-19 is associated with sociodemographic factors and with chronic conditions such as diabetes, asthma, circulatory disease and others. However the associations with pre-existing disease are not just with a small set of conditions that contribute to risk, but with many conditions as demonstrated by associations with past medical and prescribing history in relation to multiple physiological systems. As countries attempt to emerge from lockdown whist protecting vulnerable individuals, multivariable classifiers rather than crude rule-based approaches will be needed to define those most at risk of developing severe disease.

## Data Availability

Data cannot be directly shared publicly because of ethical and legal concerns as the data derive from deidentified National Health Service Records. The datasets used in this analysis are available via the Public Benefits Privacy Panel for Health at https://www.informationgovernance.scot.nhs.uk/pbpphsc/ for researchers who meet the criteria for access to confidential data.

## Declarations

### Information governance

This study was conducted under approvals from the Public Benefit and Privacy Panel for Health and Social Care that allow Public Health Scotland staff to link datasets. Datasets were de-identified before analysis.

### Data availability

The component datasets used here are available via the Public Benefits Privacy Panel for Health at https://www.informationgovernance.scot.nhs.uk/pbpphsc/ for researchers who meet the criteria for access to confidential data. All source code used for derivation of variables, statistical analysis and generation of this manuscript is available on https://github.com/pmckeigue/covid-scotland_public

## Acknowledgements

We thank all staff in critical care units who submitted data to the SICSAG database, the Scottish Morbidity Record Data Team, the staff of the National Register of Scotland, the Public Health Scotland Terminology Services, the HPS COVID-19 Laboratory & Testing cell and the NHS Scotland Diagnostic Virology Laboratories, and Nicola Rowan (HPS) for coordinating this collaboration.

## Public Health Scotland COVID-19 Health Protection Study Group

Alice Whettlock^1^, Allan McLeod^1^, Andrew Gasiorowski^1^, Andrew Merrick^1^, Andy McAuley^1^, April Went^1^, Calum Purdie^1^, Colin Fischbacher^1^, Colin Ramsay^1^, David Bailey^1^, David Henderson^1^, Diogo Marques^1^, Eisin McDonald^1^, Genna Drennan^1^, Graeme Gowans^1^, Graeme Reid^1^, Heather Murdoch^1^, Jade Carruthers^1^, Janet Fleming^1^, Jade Carruthers^1^, Joseph Jasperse^1^, Josie Murray^1^, Karen Heatlie^1^, Lindsay Mathie^1^, Lorraine Donaldson^1^, Martin Paton^1^, Martin Reid^1^, Melissa Llano^1^, Michelle Murphy-Hall^1^, Paul Smith^1^, Ros Hall^1^, Ross Cameron^1^, Susan Brownlie^1^, Adam Gaffney^2^, Aynsley Milne^2^, Christopher Sullivan^2^, Edward McArdle^2^, Elaine Glass^2^, Johanna Young^2^, William Malcolm^2^, Jodie McCoubrey ^2^

1 Health Protection Scotland (Public Health Scotland), Meridian Court, 5 Cadogan Street, Glasgow G2 6QE.

2 NHS National Services Scotland, Meridian Court, 5 Cadogan Street, Glasgow G2 6QE.

## Supplementary tables

**Table S1.**
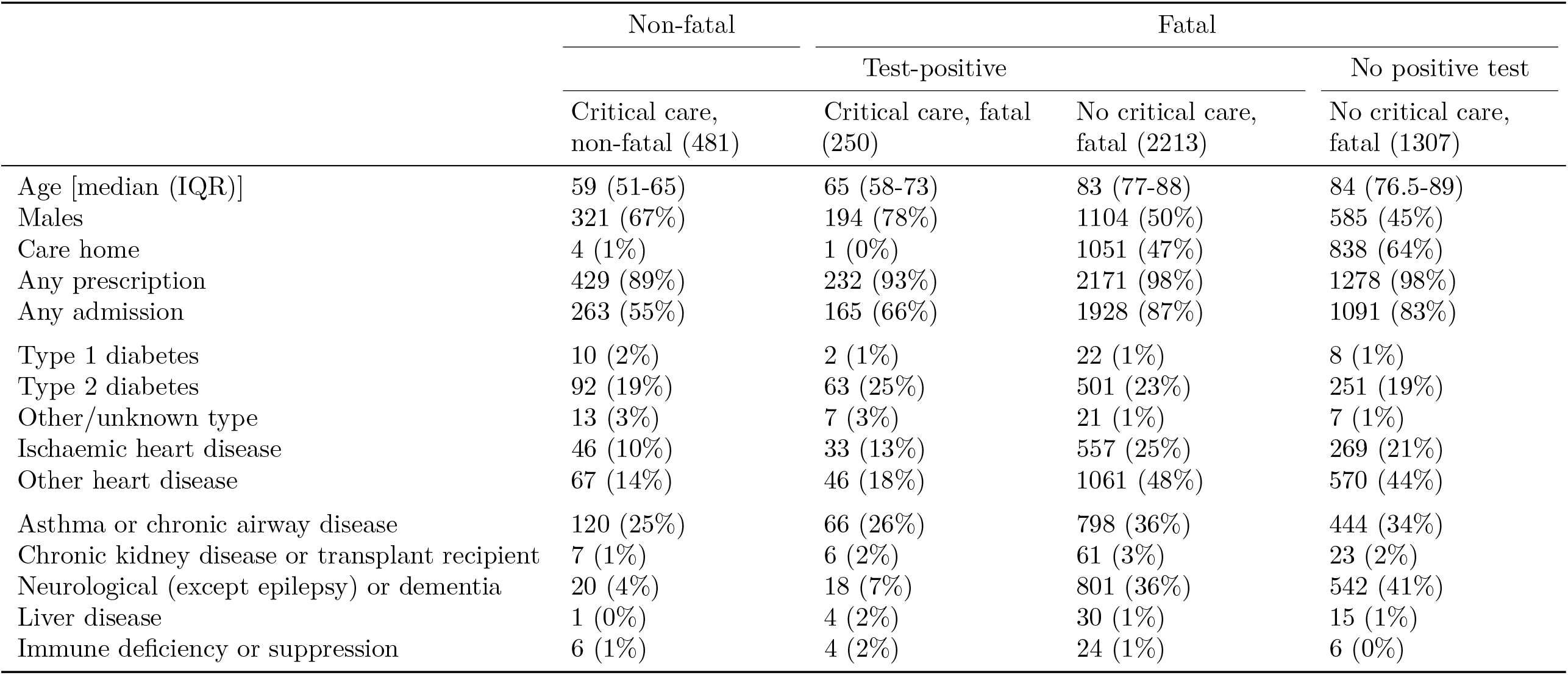
Comparison of severe non-fatal and fatal cases, by test positive status and entry to critical care

**Table S2.**
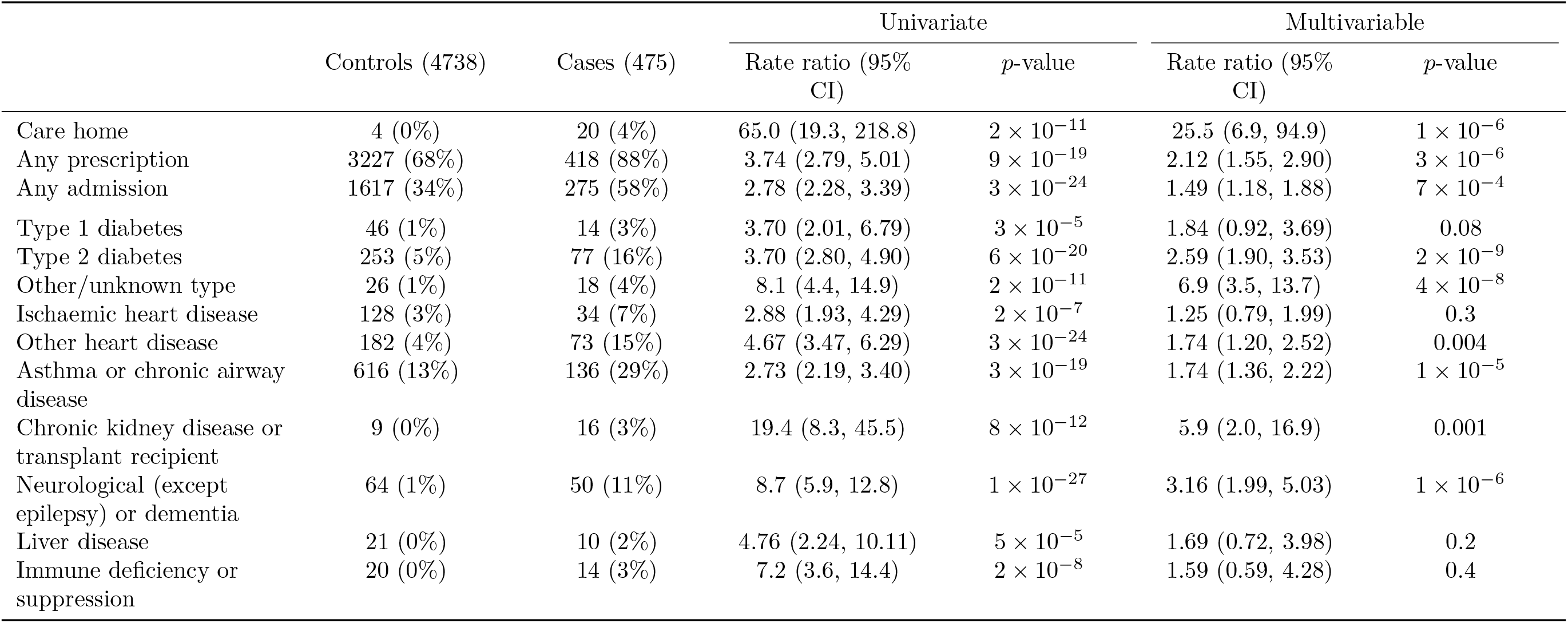
Associations of severe disease with listed conditions in those aged less than 60

**Table S3.**
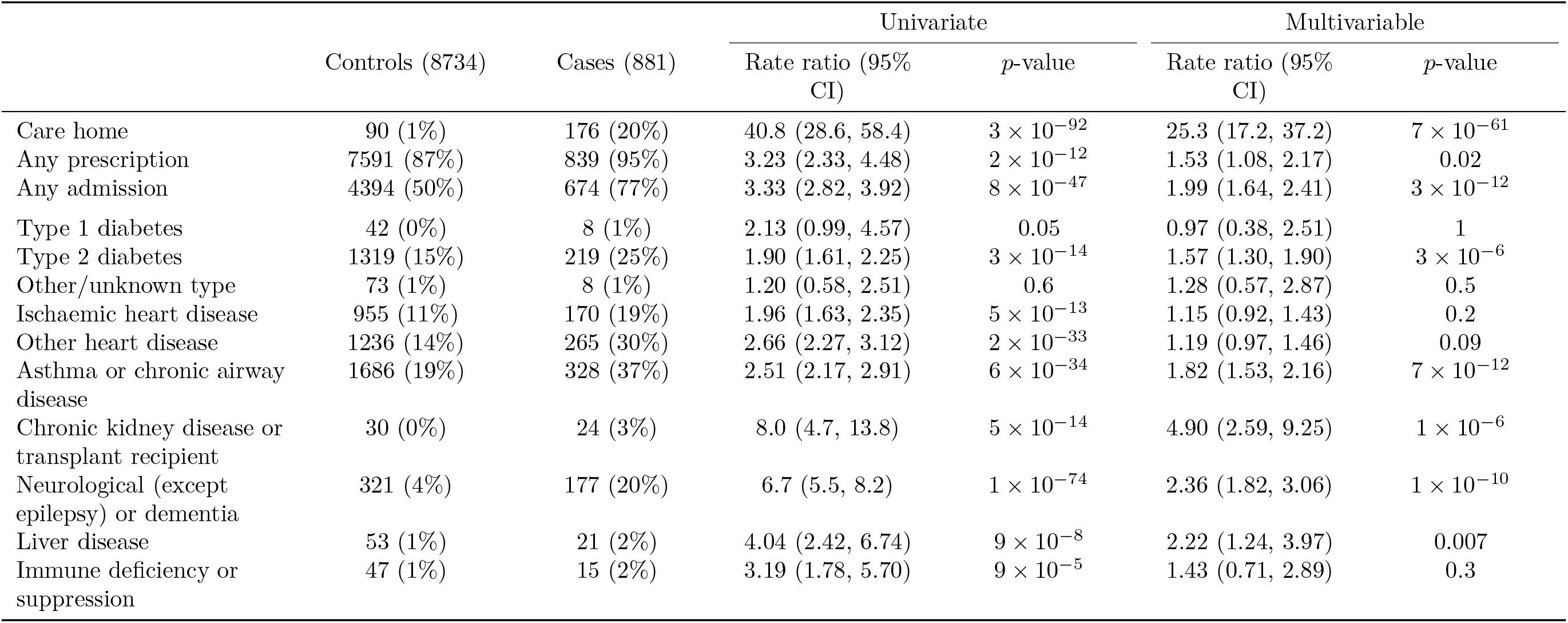
Associations of severe disease with listed conditions in those aged 60-74 years

|  |  | Controls (8734) | Cases (881) | Univariate |  | Multivariable |  |
| --- | --- | --- | --- | --- | --- | --- | --- |
|  |  |  |  | Rate ratio (95% CI) | p-value | Rate ratio (95% CI) | p-value |
| Care home | | 90 (1%) | 176 (20%) | 40.8 (28.6, 58.4) | $3 \times 10^{-92}$ | 25.3 (17.2, 37.2) | $7 \times 10^{-61}$ |
| Any prescription | | 7591 (87%) | 839 (95%) | 3.23 (2.33, 4.48) | $2 \times 10^{-12}$ | 1.53 (1.08, 2.17) | 0.02 |
| Any admission | | 4394 (50%) | 674 (77%) | 3.33 (2.82, 3.92) | $8 \times 10^{-47}$ | 1.99 (1.64, 2.41) | $3 \times 10^{-12}$ |
| Type 1 diabetes |  | 42 (0%) | 8 (1%) | 2.13 (0.99, 4.57) | 0.05 | 0.97 (0.38, 2.51) | 1 |
| Type 2 diabetes | | 1319 (15%) | 219 (25%) | 1.90 (1.61, 2.25) | $3 \times 10^{-14}$ | 1.57 (1.30, 1.90) | $3 \times 10^{-6}$ |
| Other/unknown type |  | 73 (1%) | 8 (1%) | 1.20 (0.58, 2.51) | 0.6 | 1.28 (0.57, 2.87) | 0.5 |
| Ischaemic heart disease | | 955 (11%) | 170 (19%) | 1.96 (1.63, 2.35) | $5 \times 10^{-13}$ | 1.15 (0.92, 1.43) | 0.2 |
| Other heart disease | | 1236 (14%) | 265 (30%) | 2.66 (2.27, 3.12) | $2 \times 10^{-33}$ | 1.19 (0.97, 1.46) | 0.09 |
| Asthma or chronic airway disease | | 1686 (19%) | 328 (37%) | 2.51 (2.17, 2.91) | $6 \times 10^{-34}$ | 1.82 (1.53, 2.16) | $7 \times 10^{-12}$ |
| Chronic kidney disease or transplant recipient | | 30 (0%) | 24 (3%) | 8.0 (4.7, 13.8) | $5 \times 10^{-14}$ | 4.90 (2.59, 9.25) | $1 \times 10^{-6}$ |
| Neurological (except epilepsy) or dementia | | 321 (4%) | 177 (20%) | 6.7 (5.5, 8.2) | $1 \times 10^{-74}$ | 2.36 (1.82, 3.06) | $1 \times 10^{-10}$ |
| Liver disease | | 53 (1%) | 21 (2%) | 4.04 (2.42, 6.74) | $9 \times 10^{-8}$ | 2.22 (1.24, 3.97) | 0.007 |
| Immune deficiency or suppression | | 47 (1%) | 15 (2%) | 3.19 (1.78, 5.70) | $9 \times 10^{-5}$ | 1.43 (0.71, 2.89) | 0.3 |

**Table S4.**
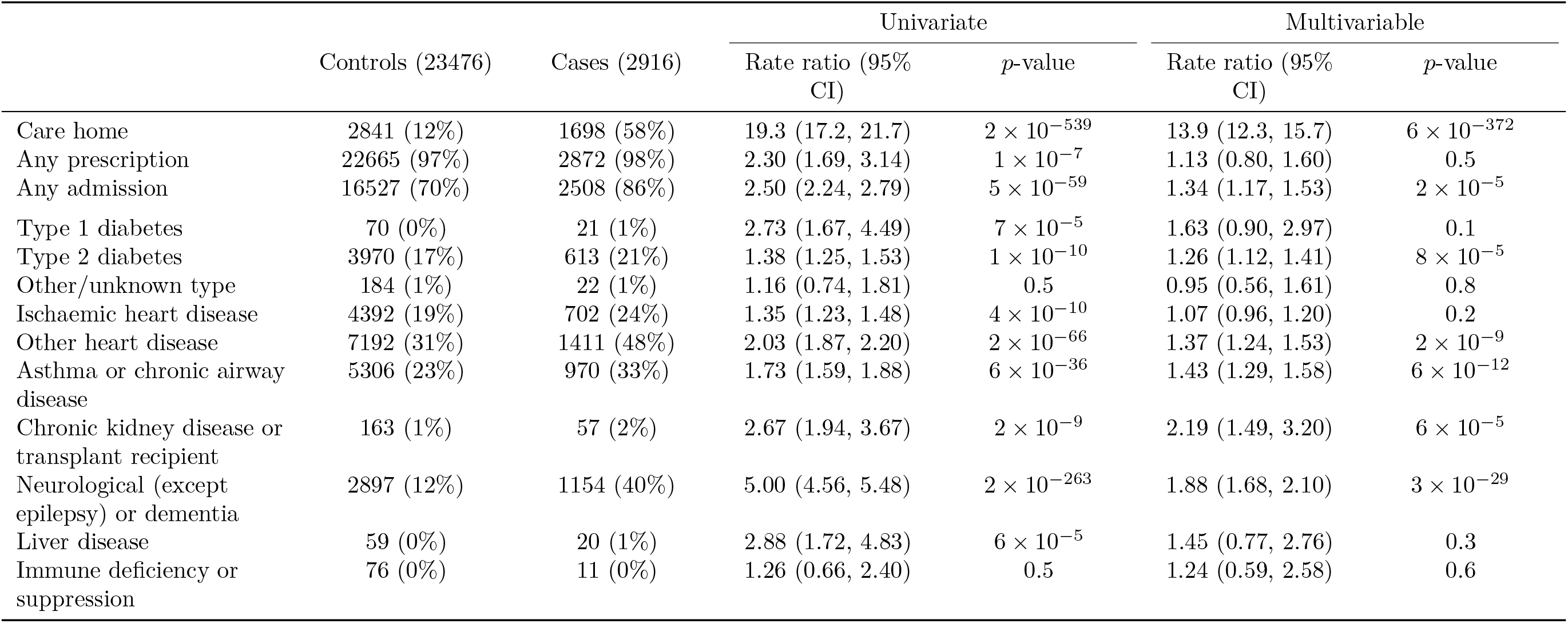
Associations of severe disease with listed conditions in those aged 75 years and over

**Table S5.**
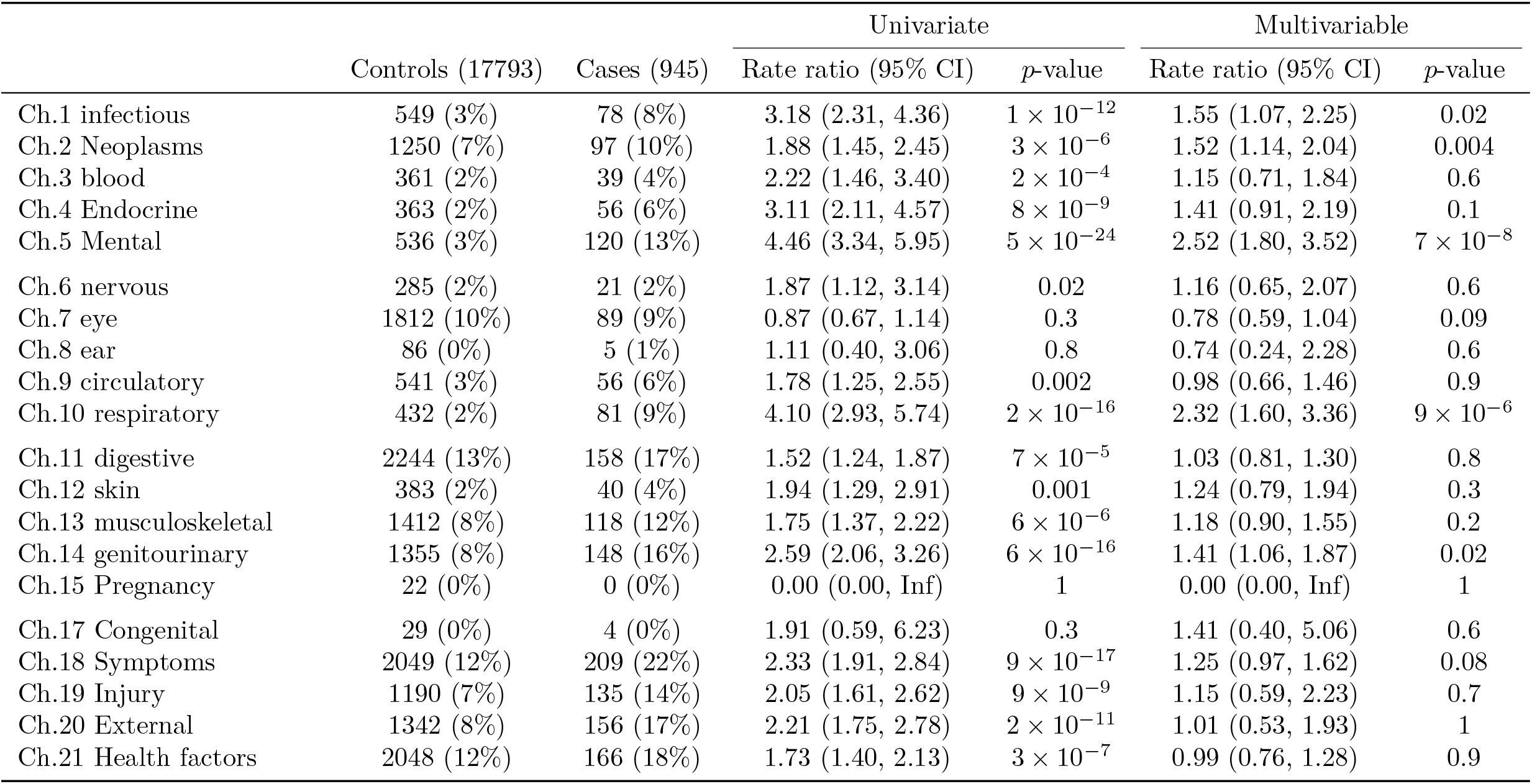
Associations of severe disease with hospital diagnoses in last 5 years, in those without any listed condition

**Table S6.**
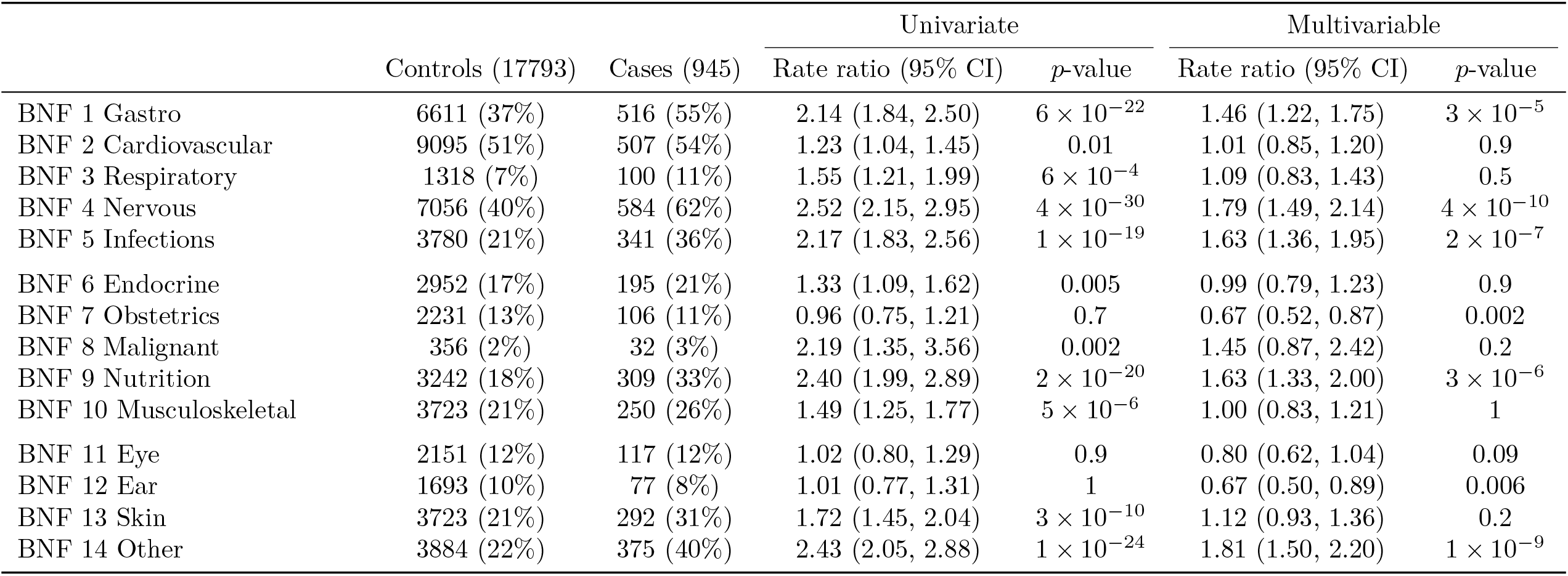
Associations of severe disease with prescribed drugs in those without any listed condition

**Table S7.**
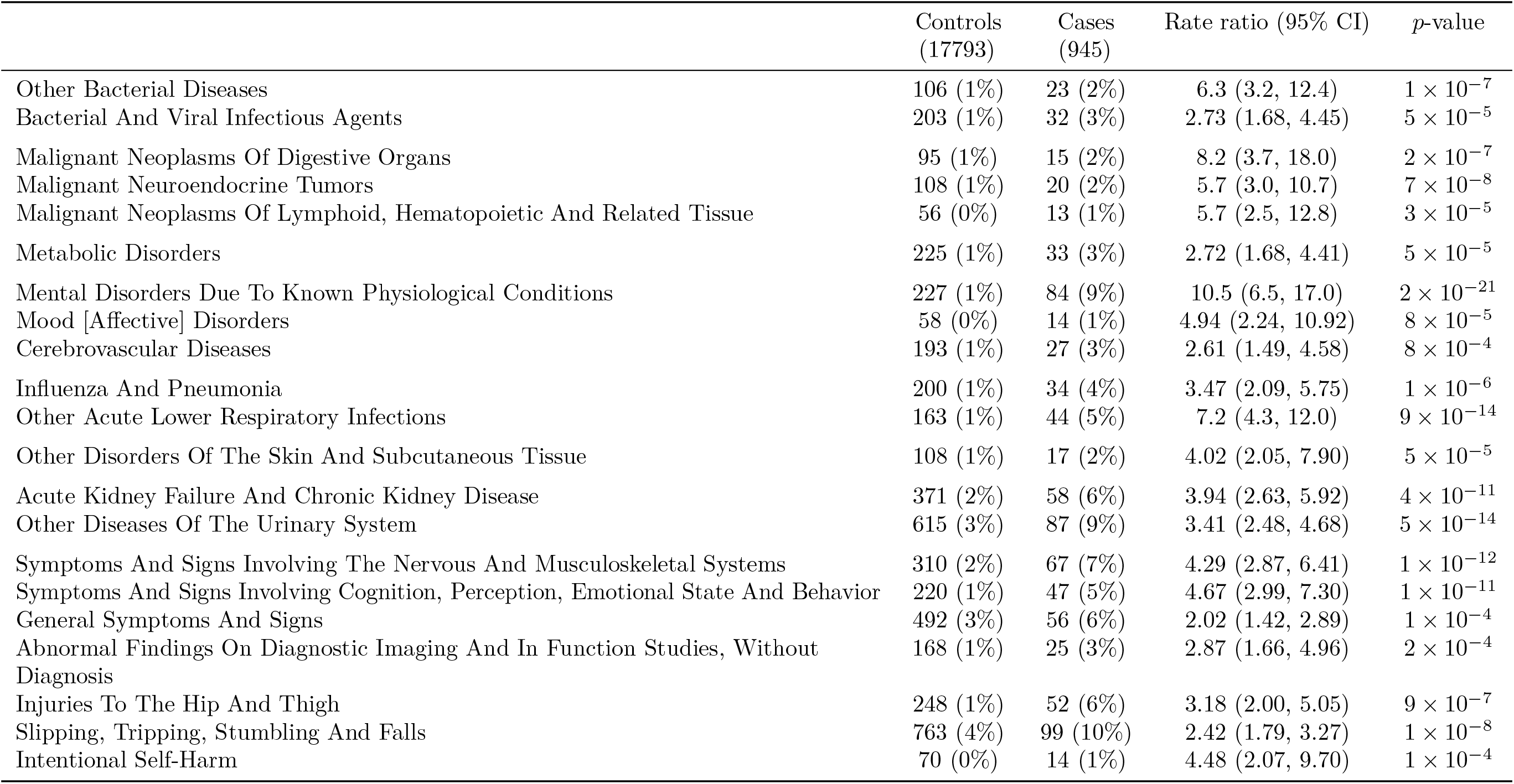
Univariate associations of severe disease with hospital diagnoses by ICD subchapters in those without any listed conditions: rows retained are those with p < 0.001 and at least 50 cases and controls Symptoms And Signs Involving Cognition, Perception, Emotional State And Behavior 220 (1%)

**Table S8.**
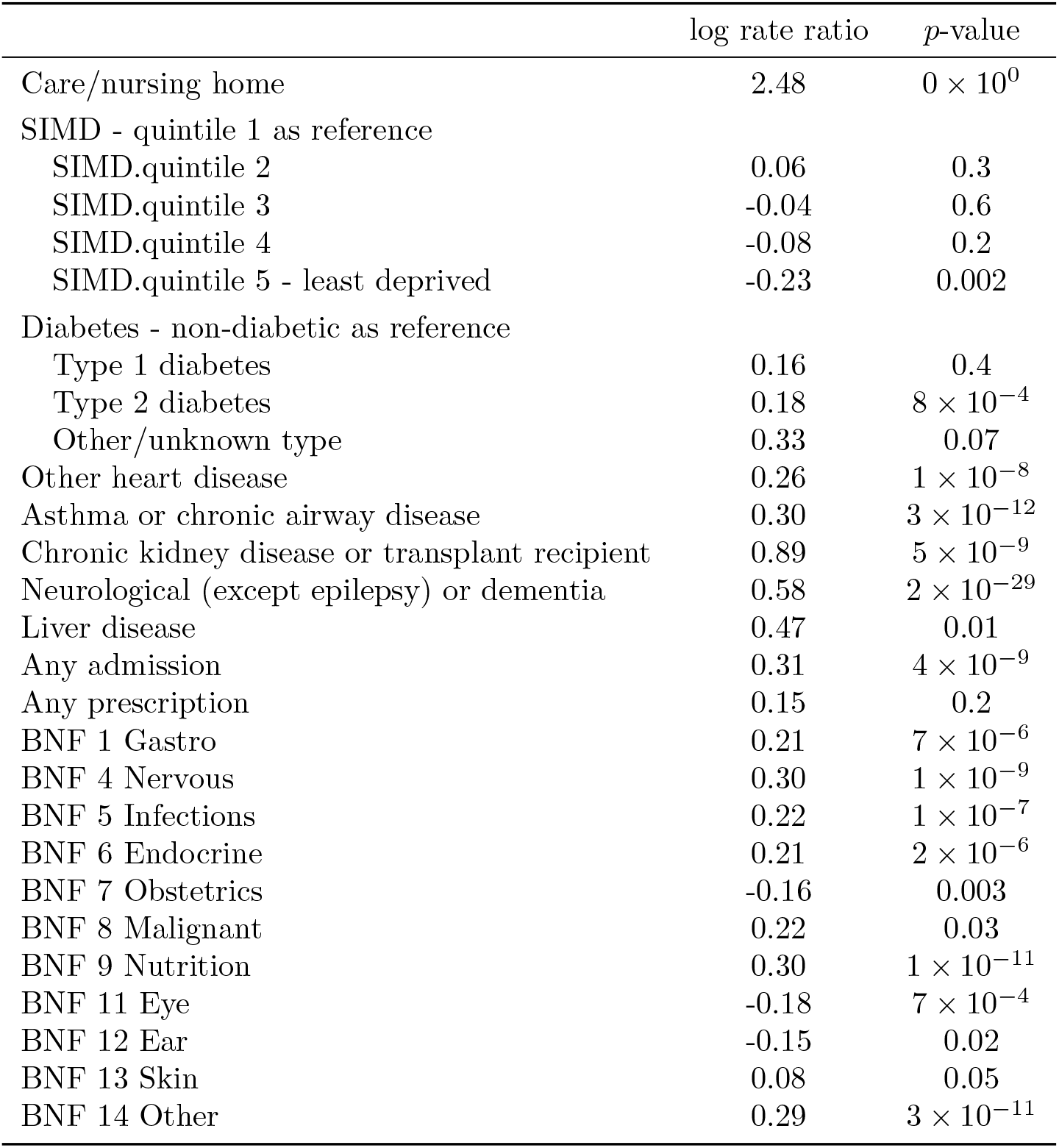
Stepwise regression: variables retained in model for severe disease

